# Face Identity Recognition with Interference of Unusual Features by People with Schizophrenia

**DOI:** 10.64898/2026.02.07.26345453

**Authors:** Melina Medeiros de Miranda Lima, Aline Mendes Lacerda, Maria Lúcia de Bustamante Simas, Nelson Torro Alves

## Abstract

Schizophrenia is a serious mental disorder characterized by enhanced sensory-perceptual alterations. We investigated face identity recognition in people with schizophrenia with the Facial Identity Recognition Structured Task (FIRST) develop at our laboratory. This was created with natural interference features (beard, makeup and mask). This task consists in six block-trails of six images for identity recognition. Forty three adult volunteers divided into two groups: a Health Control (HC) and a group of hospitalized patients with Schizophrenia (SchG) participated in the study. We measured the total number of correct answers as well as the average reaction time for each block. We observed significant losses in recognition of identity faces with interferences such as make up, beard and facial-mask.

## Introduction

Schizophrenia is a severe neuropsychiatric disorder involving multidimensional impairments in sensation and perception (de Bustamante Simas et al., 2021, Silverstein, Demmin, & Bed-nar, 2017), reasoning and thinking (Palominos, Figueroa-Barra, & Hinzen, 2023), emotion (Gong, Li, Zhao, &Wu, 2021), and behavior (Peng, Li, Lv, Zhang, & Zhan, 2018), associated to a mild or severe cognitive impairment (Silberstein, & Harvey, 2019). These dysfunctions can impact social cognition in part caused by deficits in facial processing (Marwick, & Hall, 2008, Guillaume, & Thomas, 2021, Liu *et al*., 2021, Yang, *et al*., 2018, Darke, Sundram, Cropper, & Carter, 2021, Ohara, *et al*., 2020, Ferroni, *et al*., 2019

Ohara et al. (2020), by using magnetoencephalography (MEG), compared the response of patients with schizophrenia and healthy controls to human faces and objects. They found specific reductions in M170 to faces only for patients with schizophrenia. Chen and Ekstrom (2016) compared recognition of faces using a procedure that involved qualitative and quantitative judgements. A self-report questionnaire evaluated the subjective perception of faces, while an objective paradigm measured thresholds to discriminate face identities. The task involved recognizing a target face from two face stimuli in the sequence. Patients with schizophrenia showed worse performance in the objective assessment as well as in the self-report, but these two measures were not correlated. The authors concluded that the functional relationship between face processing and everyday subjective experience is impaired in schizophrenia.

However, most of the studies have focused on emotional expressions (Yang et al., 2018), memory components (Guillaume, & Thomas, 2021) or used computation resources and artificial facial characteristics (Chen, & Ekstrom, 2016). Differently, in the present work, we investigated the simultaneous perception of neutral faces by presenting more ecological stimulus (moving faces). In the task, participants had to recognize people with and without beard, with and without make-up, and with and without facial-mask, considered as features that disturbs the perception (FDP).

The task involved the identification of a target face presented simultaneously to the distractor faces. In five block-trials, ecological movements were used as well manipulated natural face characteristics such as beard and make-up. Additionally, in the context of the pandemics, masks were included to evaluate this unusual interference in face detection.

This procedure was based on the clinical complaints of patients with schizophrenia about the difficulties in recognizing faces during psychotic breaks in daily life. Our psychological instrument, was named Face Identification Recognition Structured Task (FIRST).

## Method

### Participants

Twenty-three volunteers from the Hospital Ulysses Pernambucano took part in the study. One volunteer did not complete the task and was excluded. Therefore, the clinical sample comprised 22 patients (15 men and 07 women) with schizophrenia (SchG). The Healthy Control Group (HC) included 20 volunteers (12 men and 08 women).

The experiments were run after approval by the ethics committee from CCS UFPE (CAAE: #57285022.0.0000.5208, review #5.414.053).

### Equipment and Instruments

#### Screening assessment

(i) visual acuity test for cellphones with optotypes “E” from Rasquin (*Smart Optometry* IOS app developed by Smart Optometry Ltd.); (ii) sociodemographic, familial and clinical background interview; (iii) Mini Mental State Examination (MMSE).

#### Face Identity Recognition Structured Task (FIRST) design

Face Identity Recognition Structured Task (FIRST) was designed and developed at and by the Laboratório de Percepção Visual from the Universidade Federal de Pernambuco (LabVis – UFPE) with the objective of testing the difficulties of people with schizophrenia in identifying people in conditions yet to be tested.

In FIRST all possible responses were presented in static and grayscale mode. In block-trial 1, male and female target faces were frontal and statical as well as the responses. In block-trials 2 to 6, male and female target faces were movie shots in color and in natural 180° horizontal motion. In block-trial 2 the possible responses were only frontal. In block-trial 3 the possible responses were faces at 45º. In block-trial 4, possible responses were frontal faces with makeup (women) or beard (men). Block-trial 5 was similar to block-trial 4, but possible responses were faces at 45º. Finally, in block-trial 6, the possible responses wearing (covid) facial mask.

The first laminae of block-trials 1 and 2 are training examples. Responses were recorded by the experimenter who made a log in the Timestamper app at the end of each response.

#### Data processing software

(i) IOS app *Timestamper: Keep Activity Log* by Susamp infotec; (ii) *Statistica* 14.0 by Tibco and (iii) *GraphPad Prism* 9 by Dotmatics

#### Equipment

(i) 15” Notebook L12D1DJ2, Intel(R) Core(TM) i5-1035G1, 15’’, (ii) iPhone 11, iOS 15.4

### Procedure

We started the experimental session with the interview followed by the visual acuity test and the MMSE. Only then we run the main experiment.

The Face Identity Recognition Structured Task (FIRST) composed of six block-trials of increasing difficulty, each block consisting in six laminae, was presented in a 15-inch notebook display at an average distance of about 57 centimeters. The given instructions were: “a face will be presented, and you must say which one of the faces at the bottom corresponds to the top stimulus face. In other words, you must select among the faces at the bottom whose identity coincides with the person in the top stimulus. Please, give your response as quickly as you can by naming the number of the selected face, i.e. 1, 2, 3 or 4.”. The whole experiment was run in a single session, lasting about 50 minutes.

### Data Analysis

Age and responses from the interviews and MMSE were tabled in a single sheet (see Table 1). Non-parametric Mann Witney U Tests were run to evaluate age and MMSE differences between samples (SchG and HC). Visual acuity equal to 1 was used as an exclusion criterion.

Tabled results from FIRST were organized into number of correct responses (CR) and reaction time (RT). Sum, mean and standard deviation of CR were estimated across participants from HC and SchG. On the other hand, only mean and standard deviation across participants were taken as estimates for RT from both groups. To test differences between groups we used the Mann Whitney U Test, and for differences between block-trials the Wilcoxon Test. Considering the number of simultaneous comparisons (six), we used Bonferroni correction (p<0.008).

We also performed correlation tests (Spearman Coefficient) between CR and RT results and MMSE, education and self-rating values (0 – 4) for easiness of face recognition. We once again used the Bonferroni correction to consider significant correlations.

## Results

Our findings shows SchG slightly under the average as compared to HC, but this self-rated difference did not reach significance. Nevertheless, rates for MMSE, as expected, showed lower cognitive performance for the SchG. Table 1 also reports ratings, from 0 (very bad) to 4 (very good), given by the volunteers about their own ability to recognize faces.

Perfect educational level matching was not possible, but the difference between mean age was found to be not significant.

Figure 1 shows CR for SchG and HC. The SchG had fewer CR in all block-trials. There were significant differences in all block-trials, except for number two.

**Figure 1.**
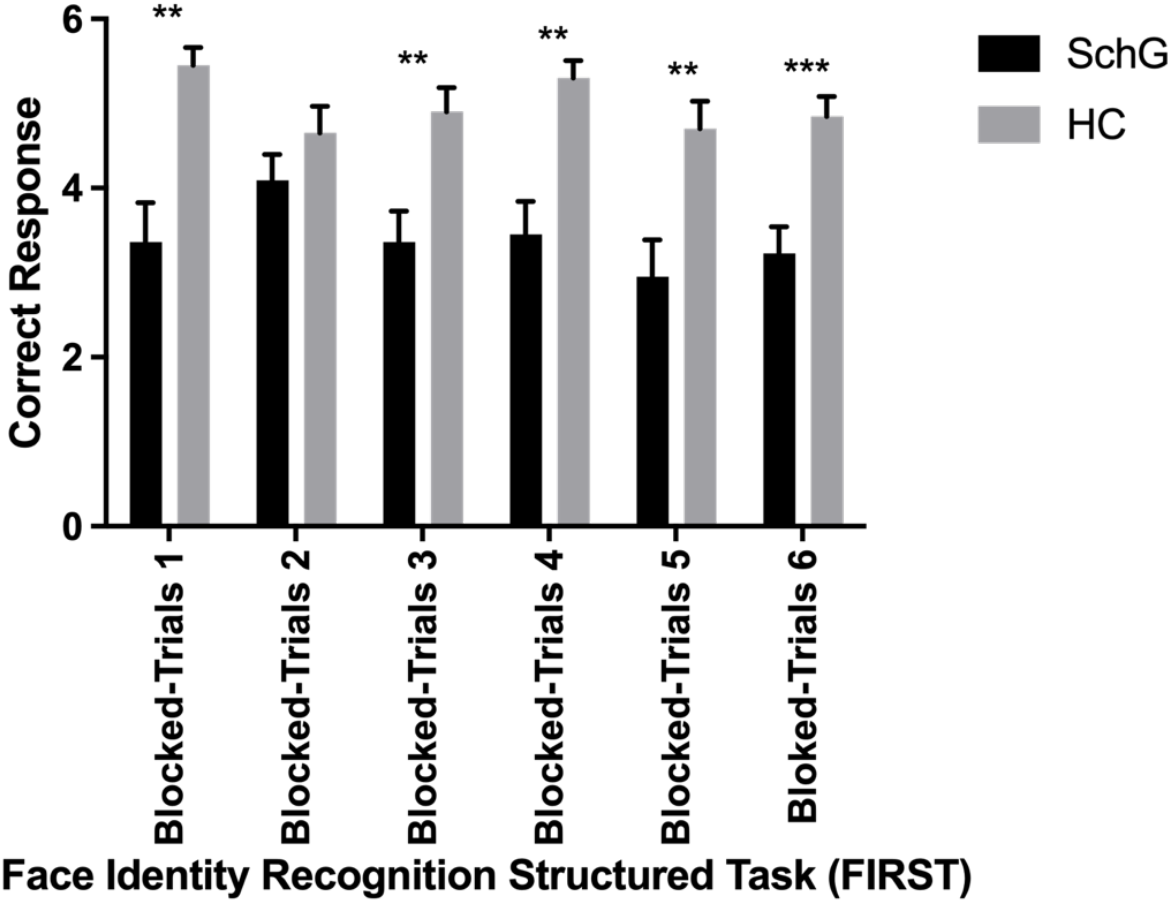
Mean CR per group per block-trial. Note. Mann Whitney U Test: *p > .05; **p > .001; *** p > .0001.

**Figure 2.**
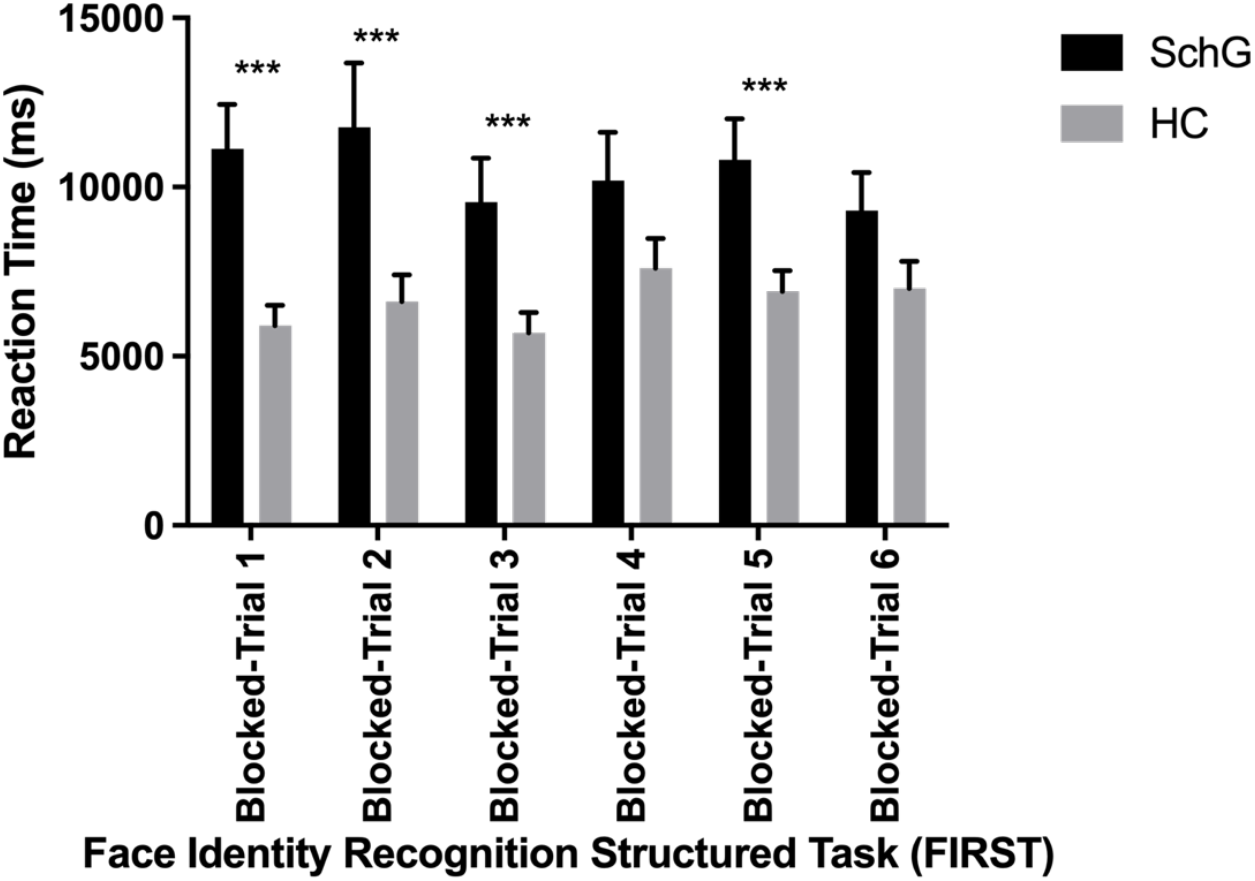
Mean RT per group per block-trial. Note. Mann Whitney U Test: *p > .05; **p > .001; *** p > .0001; BT1 (B&W frontal face, B&W frontal response); BT2 (moving color face, B&W frontal response); BT3 (moving color face, B&W 45º face response); BT4 (moving color face, B&W frontal face with make up or beard); BT5 (moving color face, B&W 45º face with make up or beard); BT6 (moving color face, B&W covid mask).

The easiest block-trial was the first followed by, the fifth, the fourth, the sixth, the third and the second.

Reaction Time starting from the onset of the stimulus and ending with the response by the participant was recorded for each lamina. Only then the mean RT was taken for each block-trial. Mean RT was always greater for SchG than HC, and significant differences were found in four of the six block-trials. Exceptions were block-trials two and six. Reaction time were higher for the SchG and ranged between 2.0 and 1.33 fold slower than HC.

Figure 3 shows SchG/HC ratio between mean CRs and mean RTs for each BT. When evaluating the effect of intervening features by direct comparison of ratio mean from BT2 to that for BT4, and from BT3 to that for BT5, we did find that make up (in women) and beard (in men) indeed showed a raise of response error in SchG as compared to HC. However, this difference did not achieve significance. Neither did any other comparison between BTs (p < 0.008), nor for each group nor for the entire sample.

**Figure 3.**
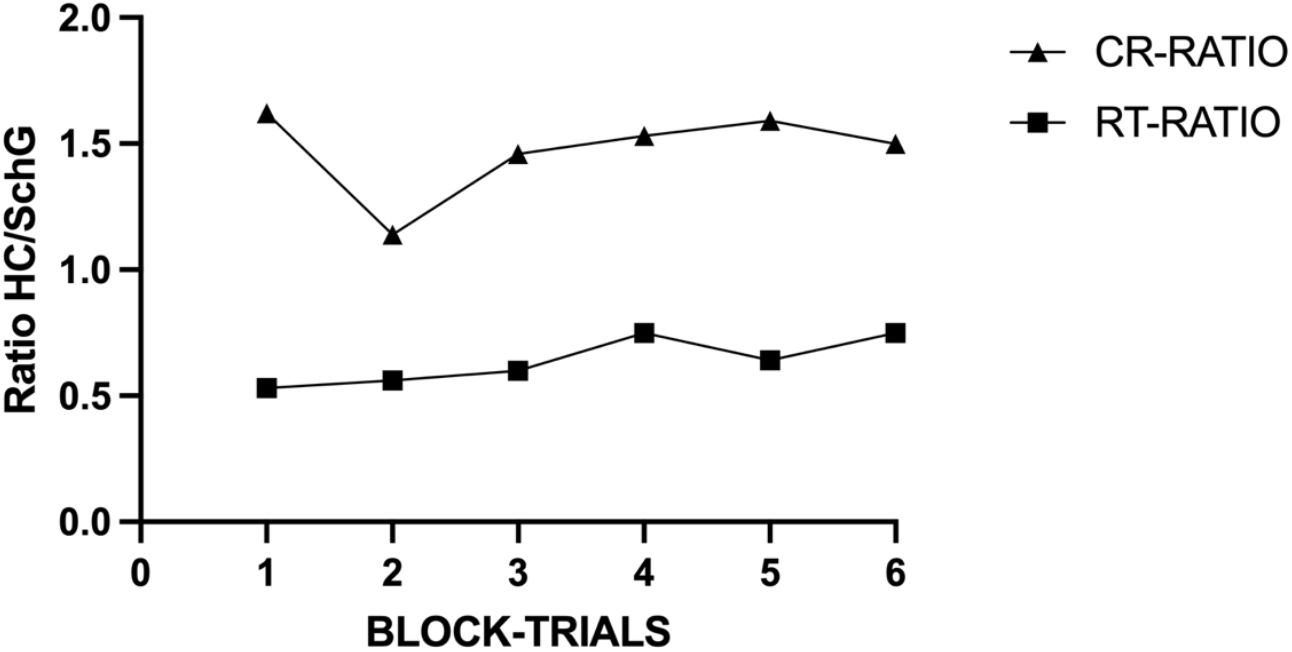
CR and RT ratio of mean for HC and SchG.

We also found correlations among the data. A positive correlation (r_s_ = 0.52, p < .008, N = 43) between education and MMSE, and of education and CR (r_s_ = 0.56, p < .008, N = 43). The MMSE also showed strong and positive correlation with CR (r_s_ = 0.71, p < .008, N = 43). On the other hand, there was a moderate negative correlation between education and RT(rs = -0.41, p < 0.008, N = 43), and a strong negative correlation between MMSE and RT (r_s_ = -0.60, p < .008, N = 43). We also found some weak and positive correlation between self-rating of easiness of face recognition and another variable, mostly CR or RT.

There were no significant correlations among variables within each group.

## Discussion

Our findings coincide with those of the literature (Guillaume, & Thomas, 2021, Liu, *et al*., 2021, Yang, *et al*., 2018, Darke, Sundram, Cropper, & Carter, 2021, Ohara, *et al*., 2020, Ferroni, *et al*., 2019) that report losses in identity face recognition in people with schizophrenia. We believe this difficulty is related to deficits in spatial frequency contrast sensitivity (Fernandes, Shaqiri, Brand, Nogueira, Herzog, Roinishvili, Santos, & Chkonia, 2019) as well as to dysfunctional lateral inhibition, implying in difficulties of edge detection. Further, we can also take into consideration the dysfunctional saccadic eye-movements (Myles, Rossell, Phillipou, Thomas, & Gurvich, 2017), together with frontal eye-field processing difficulties (Mueller, Krock, Shepard, & Moore, 2020). The involvement of dopaminergic unbalance affecting retinal ganglion cell functions and processing must also be considered (Bodis-Wollner, 1990).

We did observe that SchG had difficulty processing faces with make-up or beard as well (BT4 and BT5). We think this is somewhat related to losses in high spatial frequency contrast sensitivity in schizophrenia (Laprevote, Oliva, Ternois, Schwan, Thomas, & Boucart, 2013) as well as lateral inhibition and dopaminergic retinal unbalance.

Another point to be raised is that most people with schizophrenia do not realize their difficulty in recognizing faces (Chen and Ekstrom, 2016), and FIRST was designed exactly to investigate this difficulty mentioned by some patients within the clinical setting.

We also found some expected correlations between group sample characteristics and CR, and RT, showing that higher educational level and better cognitive performance were directly related to increase in CR and decrease in RT.

Our main finding, nevertheless, was to measure the difficulty of people with schizophrenia to recognize face identity even in the presence of the face itself, that is, without the memory component. We did introduce features like beard and make-up that interfered with the recognition process and did make difficult to identify the person despite its simultaneous presence in the display. We also introduced movement as an ecological factor assuming that face recognition is worse under steady and fixed conditions. We believed that it would facilitate identity recognition and assume that it probably did increase CR levels in SchG. This fact becomes more evident in BT2 where SchG did not show significant difference from HC. Observe that SchG did improve from BT1 to BT2 with the introduction of movement. However, it did not help improve CR ratings of SchG as compared to HC when the response involved 45º, beard, makeup or Covid mask. Unfortunately, we could not test every desired condition for comparisons purpose due to the length of the experiment.

## Conclusions

In sum, these findings corroborate our hypothesis that the loss of identity recognition also suffers interference from natural features.

The present study introduced FIRST, an identity recognition test for faces that does not involve the memory component. We did not use emotional expressions or artificial manipulation of facial characteristics, but natural features like beard and everyday make-up. Additionally, we used Covid masks.

This study presents a strong contribution to clinical and psychotherapy settings that involve schizophrenia patients. We believe psychoeducation is a crucial tool to improve quality of life in schizophrenia. Learning about alterations that occur in sensory perception in schizophrenia can help patients to deal with their own misperceptions as well as to better evaluate their own state of mind, and to make known the help they need when they need it.

Our study does have some limitations being the main one the very restricted number of faces in our bank. To have men growing their natural beard and be ready to shave them on demand is difficult. We did have some friends doing so. Another slight limitation is that RTs were measured by hand using an Android/IOS app.

## Data Availability

All data produced in the present study are available upon reasonable request to the authors

https://www.dropbox.com/scl/fi/g8wy27ajoehrtbn2a1e4t/Manuscrito-final.pdf?rlkey=p23n78jtaweal0ktvookv55s8&st=pjknu6c1&dl=0

